# Translating CRISPR-Cas Systems into musculoskeletal medicine and orthopaedics: a scoping review protocol

**DOI:** 10.1101/2025.09.14.25335710

**Authors:** Oladapo Ekundayo, Samuelson E. Osifo, Mutaleeb A. Shobode

**Author notes:** Corresponding author (OE). These authors contributed equally to this work (Oladapo Ekundayo, Samuelson E. Osifo).

## Abstract

**Background:** Musculoskeletal diseases are a leading cause of global disability and healthcare burden, yet traditional orthopaedic procedures often fail to address molecular drivers of degenerative and genetic conditions. CRISPR-Cas Systems, a precise and programmable genome-editing technology, show promise in preclinical musculoskeletal models, ranging from gene knockouts in osteoarthritis to mutation correction in skeletal dysplasia. Despite growing interest, no comprehensive synthesis exists to map how CRISPR-Cas Systems are being applied in orthopaedic research. This scoping review aims to fill that gap.

**Methods:** This protocol will be registered with the Open Science Framework and follows PRISMA-ScR guidelines and the Arksey & O’Malley framework. MEDLINE, Embase, Scopus, Web of Science, and grey literature sources from 2005 onward will be searched. Inclusion criteria encompass original research using CRISPR-Cas Systems in human, animal, or in vitro musculoskeletal models. Two reviewers will independently screen titles, abstracts, and full texts using Covidence software. Data extraction will be standardized and performed in duplicate. Extracted variables include study design, model system, target tissue, gene-editing approach, delivery system, and reported outcomes. Results will be synthesized descriptively and thematically.

**Expected Results:** We anticipate mapping the evolution and diversity of CRISPR-Cas Systems applications across musculoskeletal tissues (bone, cartilage, tendon, muscle), highlighting domains such as tissue regeneration, gene correction, and disease modeling. Delivery strategies (viral vectors, nanoparticles) and translational challenges, including off-target effects and delivery barriers, will be summarized.

**Conclusions:** This will be the first scoping review to systematically characterize the role of CRISPR-Cas Systems in musculoskeletal medicine. Findings will inform researchers, clinicians, and policymakers, helping to guide future translational research and accelerating the integration of gene editing into clinical practice.

## Introduction

The advent of Clustered Regularly Interspaced Short Palindromic Repeats (CRISPR) and associated Cas endonuclease technology represents a revolutionary development in molecular biology and regenerative medicine. Initially discovered as part of a bacterial adaptive immune response, the CRISPR-Cas Systems has been harnessed for precise genome editing, allowing site-specific double-stranded DNA cleavage under the guidance of a synthetic single-guide RNA (sgRNA)(1). This programmable tool has rapidly transformed basic and translational science, enabling efficient and cost-effective manipulation of genomic sequences in eukaryotic cells. Its broad utility in disease modeling, gene correction, and tissue engineering has catalyzed novel therapeutic strategies across a range of medical disciplines, including oncology, neurology, hematology, and increasingly, orthopaedics and musculoskeletal (MSK) medicine(2).

Musculoskeletal diseases constitute a major global burden of disease, accounting for substantial morbidity, chronic disability, and healthcare expenditure. Disorders such as osteoarthritis, intervertebral disc degeneration, muscular dystrophies, and rare skeletal dysplasias are characterized by complex pathogenesis involving genetic predispositions, mechanical stressors, inflammatory cascades, and impaired tissue regeneration(3,4). Conventional orthopedic therapies—ranging from pharmacological symptom management to surgical joint reconstruction—are often limited by their inability to address the molecular underpinnings of disease progression. Consequently, there is an increasing need for biologically driven interventions that can modify disease trajectories, enhance tissue repair, and potentially provide curative outcomes.

In this context, CRISPR-Cas gene editing offers a paradigm shift. Preclinical studies have demonstrated the successful use of CRISPR to knock out deleterious genes such as matrix metalloproteinase-13 (MMP-13), a catabolic enzyme implicated in cartilage breakdown in osteoarthritis(5); to upregulate regenerative transcription factors like SOX9 for enhanced chondrogenesis(6); and to model rare genetic skeletal disorders such as osteogenesis imperfecta through targeted mutation of COL1A1 and COL1A2 genes(7). The technology has also been applied to enhance the biological performance of mesenchymal stem cells (MSCs), either by silencing senescence pathways (e.g., p16INK4a) or promoting lineage-specific differentiation(8). These advances underscore the potential of CRISPR-Cas9 not only as a tool for understanding musculoskeletal pathophysiology, but also as a therapeutic modality to regenerate bone, cartilage, muscle, and tendon tissues.

Recent years have also witnessed efforts to integrate CRISPR-Cas with biomaterials science and 3D bioprinting. Gene-edited cells seeded into scaffolds or hydrogels are being explored to create bioengineered constructs capable of structural and functional joint restoration(9,10). Furthermore, nanotechnology-enabled delivery systems—such as lipid nanoparticles and viral vectors—have been developed to enhance the tissue specificity and efficiency of CRISPR delivery in vivo, including within load-bearing musculoskeletal environments(11). These multi-modal strategies represent the frontier of regenerative orthopaedics, with implications for both acute repair and chronic disease modification.

However, the translation of CRISPR-Cas Systems into clinical musculoskeletal care is still in its infancy. Despite promising results from ex vivo and small-animal studies, challenges remain regarding off-target effects, immunogenicity of Cas proteins, and efficient delivery to avascular tissues such as cartilage. There is also limited standardization of experimental protocols, variable outcome reporting, and a lack of longitudinal data on the safety and durability of gene edits. Moreover, ethical concerns surrounding germline editing and the regulatory complexity of somatic gene therapies continue to influence clinical trial readiness in this field(12).

To our knowledge, no scoping review to date has systematically mapped the application of CRISPR-Cas Systems within the domain of orthopaedics and musculoskeletal medicine. Given the exponential growth in the number of peer-reviewed publications, the interdisciplinary nature of this field, and the urgent need to identify translational opportunities, such a review is both timely and necessary.

## Rationale

The rationale for this scoping review stems from the intersection of several critical trends: first, the expanding role of molecular biology and gene editing in surgical subspecialties; secondly, the limitations of current orthopedic treatments in addressing genetic and degenerative musculoskeletal conditions; and thirdly, the translational promise of CRISPR-Cas Systems for regenerative and precision medicine. By synthesizing the current landscape of CRISPR-related research in orthopedic surgery and musculoskeletal medicine, this review will provide a foundational knowledge base for clinicians, researchers, and policymakers aiming to understand the scope, readiness, and future direction of gene editing in musculoskeletal care.

A scoping review is the most appropriate methodological approach due to the heterogeneity of study designs, gene targets, and experimental systems in the current literature. Unlike a traditional systematic review, which addresses narrowly focused clinical questions, a scoping review allows for the mapping of diverse and emerging evidence to identify research gaps, methodological patterns, and knowledge synthesis priorities. This review will be conducted in accordance with the PRISMA Extension for Scoping Reviews (PRISMA-ScR) guidelines(13).

## Objectives

The objective of this scoping review is to systematically identify and characterize the scope of literature on CRISPR-Cas gene editing applications in orthopaedics and musculoskeletal medicine. Specifically, this review aims to:

1. Map the existing peer-reviewed literature on CRISPR-Cas use in musculoskeletal biology and orthopedic surgery, including both basic science and translational studies.
2. Classify the target tissues, diseases, and gene editing strategies employed across studies, including applications to bone, cartilage, tendon, ligament, intervertebral disc, and skeletal muscle.
3. Describe the methodological characteristics of the studies, including delivery platforms (e.g., viral vectors, lipid nanoparticles), editing approaches (e.g., knockout, knock-in, base editing), and preclinical models used.
4. Summarize reported therapeutic outcomes, including histological, biomechanical, functional, and safety-related endpoints.
5. Identify current gaps in knowledge, including limitations of delivery, concerns regarding off-target effects, ethical considerations, and regulatory barriers.
6. Provide recommendations for future research, including translational pathways, clinical trial design considerations, and interdisciplinary collaboration opportunities.

## Methodology

### Title and Registration

This scoping review protocol is titled: “Translating CRISPR-Cas Systems into musculoskeletal medicine and orthopaedics: a scoping review protocol.”

This protocol will be registered with the Open Science Framework (OSF) to ensure transparency and reproducibility. The OSF registration will include details on the study rationale, objectives, methods, and planned dissemination. The registration link will be provided in the final published version of the review.

This scoping review will adhere to the PRISMA-ScR (Preferred Reporting Items for Systematic Reviews and Meta-Analyses extension for Scoping Reviews) guidelines(13) and be guided by the methodological frameworks established by Arksey and O’Malley(14) and expanded by Levac et al (15). The following methodological steps will be implemented:

### Eligibility Criteria

#### Inclusion Criteria

- All types of original research that involve application of CRISPR-Cas9 gene editing in the orthopaedic or musculoskeletal context.
- Studies involving human subjects, animal models, or in vitro cell or tissue models.
- Studies that report outcomes relevant to musculoskeletal biology or therapy (e.g., histological, molecular, biomechanical, or functional results).
- Literature published from January 1, 2005 to the date of final search execution.
- No restrictions based on language.
- Peer-reviewed publications and grey literature (e.g., conference abstracts, theses, preprints), consistent with scoping review methodology.

#### Exclusion Criteria

- Studies that do not involve CRISPR-Cas9 gene editing.
- Literature not reporting original empirical data (e.g., opinion pieces, editorials, narrative reviews).

#### Information Sources

A comprehensive search will be performed across the following electronic databases: MEDLINE (via PubMed), Embase, Scopus, and Web of Science. Grey literature sources will include ProQuest Dissertations & Theses, preprint servers (bioRxiv, medRxiv), and Google Scholar. Reference lists of all included studies will be hand-searched to identify additional relevant articles.

#### Search Strategy

A structured search strategy will be developed in collaboration with our research librarian. The search will use a combination of controlled vocabulary (e.g., MeSH and Emtree terms) and free-text keywords including but not limited to: “CRISPR”, “CRISPR-Cas”, “gene editing”, “orthopaedics”, “musculoskeletal”, “bone”, “cartilage”, “skeletal muscle”, “tendon”, and “ligament”. The full search strategy for each database will be provided in an appendix.

#### Selection Process

All search results will be imported into Covidence systematic review software (Veritas Health Innovation, Melbourne, Australia)(16) for deduplication and screening. A pilot screening of the first 50 citations will be conducted by all three reviewers (OE, SO, MS) to ensure consistency in applying the eligibility criteria. After calibration, two reviewers (OE and SO) will independently screen the remaining titles and abstracts. Articles deemed potentially eligible will undergo full-text review by both reviewers.

Discrepancies during screening or full-text review will be resolved by discussion. If consensus cannot be reached, a third reviewer (MS) will adjudicate. Reasons for exclusion at the full-text stage will be recorded. If studies are published in duplicate (e.g., conference abstract and full paper), only the most complete version will be included.

#### Data Charting Process

Data will be extracted using a standardized charting form developed a priori and refined through pilot testing. Two reviewers will independently extract data on:

- Publication details (authors, year, country)
- Study design and model (human, animal, or in vitro)
- Target tissue or disease
- CRISPR-Cas gene editing strategy (e.g., knockout, knock-in, base editing)
- Delivery system (e.g., viral vector, nanoparticle, plasmid)
- Outcomes and main findings
- Reported challenges (e.g., off-target effects, delivery issues)

Discrepancies in data extraction will be resolved by discussion and consensus. Any failure to resolve this way will be arbitrated by MS if necessary.

#### Timeline of Study

This scoping review is planned to begin October 2025, and data collection is expected to be completed by the end of October 2025. Results are expected before end of December 2025.

#### Synthesis of Results

Results will be summarized descriptively using tables and narrative synthesis. We will provide a numerical overview of study types, publication years, targeted tissues, editing approaches, and delivery platforms. Thematic analysis will be used to identify key research domains (e.g., tissue regeneration, disease modeling, therapeutic development), highlight trends, and identify knowledge gaps.

#### Ethics and Dissemination

As this review involves only secondary analysis of published literature, ethical approval is not required. Results will be disseminated through peer-reviewed publication after the study is completed.

## Discussion

This scoping review will provide a comprehensive map of existing literature on the use of CRISPR-Cas technology in orthopaedics and musculoskeletal (MSK) medicine. By synthesizing studies across basic science, translational research, and preclinical experimentation, the review will help illuminate the extent, diversity, and trajectory of gene editing applications in this field.

The findings will contribute to an improved understanding of how CRISPR-Cas Systems has been employed to address degenerative, traumatic, and genetic musculoskeletal disorders, and the delivery systems and editing strategies most frequently explored. Furthermore, by identifying both well-developed and under-represented areas, the review will inform future research directions and help guide translational strategies.

Given the rapid pace of innovation, this review may also serve as a benchmark to assess the maturity and readiness of CRISPR-based therapies for clinical application in orthopaedic practice. The integration of biomaterials, stem cells, and gene editing represents a particularly dynamic frontier with broad implications for tissue regeneration and repair.

### Strengths and Limitations

This review will be conducted using a rigorous and transparent methodological framework (PRISMA-ScR), with systematic literature searching and independent dual review of studies. It will also include grey literature to broaden scope.

However, as with all scoping reviews, we will not perform a formal risk of bias assessment or meta-analysis. The expected heterogeneity in models, targets, and methodologies may limit comparability across studies. As no formal critical appraisal will be performed, conclusions will be limited to descriptive synthesis. Additionally, while every effort will be made to include non-English literature through translation, resource constraints may affect inclusion of some articles in rare languages.

## Data Availability

Deidentified research data will be made publicly available when the study is completed and published.

